# Automated COVID-19 Detection from Chest X-Ray Images: A High Resolution Network (HRNet) Approach

**DOI:** 10.1101/2020.08.26.20182311

**Authors:** Sifat Ahmed, Tonmoy Hossain, Oishee Bintey Hoque, Sujan Sarker, Sejuti Rahman, Faisal Muhammad Shah

## Abstract

The pandemic, originated by novel coronavirus 2019 (COVID-19), continuing its devastating effect on the health, well-being, and economy of the global population. A critical step to restrain this pandemic is the early detection of COVID-19 in the human body, to constraint the exposure and control the spread of the virus. Chest X-Rays are one of the non-invasive tools to detect this disease as the manual PCR diagnosis process is quite tedious and time-consuming. In this work, we propose an automated COVID-19 classifier, utilizing available COVID and non-COVID X-Ray datasets, along with High Resolution Network (HRNet) for feature extraction embedding with the UNet for segmentation purposes. To evaluate the proposed dataset, several baseline experiments have been performed employing numerous deep learning architectures. With extensive experiment, we got 99.26% accuracy, 98.53% sensitivity, and 98.82% specificity with HRNet which surpasses the performances of the existing models. Our proposed methodology ensures unbiased high accuracy, which increases the probability of incorporating X-Ray images into the diagnosis of the disease.

## 1 Introduction

Previously specified as 2019 novel coronavirus (2019-nCOV), the Severe Acute Respiratory Syndrome Coronavirus (SARS-CoV-2) disease (COVID-19) has precipitated global outbreak as it is termed as a pandemic by World Health Organization (WHO) [1]. It is rapidly disseminating all over the world since the development of the virus in Wuhan, China, at the end of 2019 [2] [3]. Though it was reportedly covered about the linkage of the Wuhan animal market, pointing out about the animal to human transmission, further studies have suggested human to human transmission [4] through droplets and inappropriate social distancing. Nevertheless, the virus is transmitting expeditiously, the costliest Polymerase Chain Reaction (PCR) testing and inept isolating process still behind the foundation of proper treatment. For that purpose, we’ve to think of an economical and straightforward testing approach by which the examining process can escalate with the speed of transmitting to promptly identify and detach the infected person. So, it is the most decisive task to defend the proliferation as this malignant virus is continuously ravaging the world.

The convoluted PCR testing method is the only approved process of detecting the novel COVID-19 disease by WHO. For most of the country, people cannot afford this verification cost. As a result, people are dying without getting proper treatment because of the fatal virus. Moreover, the effect of this virus reflects in the major body parts including lung, heart, brain, etc. In a study published by nature, directly induced lung injury and deteriorate the respiratory system [5]. Also, there is a considerable amount of feature and observation to distinguish the infected part from a lung image. So, it will be beneficial for the society and a milestone development if we consider and establish a detection model based on the Chest X-Ray (CXR) or CT Scan images to classify the COVID-19 disease.

Researchers around the world are continuously trying to build a time-efficient and cost-effective model to identify COVID-19 disease. Investigators are adopting CXR and CT Scan images for the classification of the infected lung. There is a disparate type of AI-based architectures developed to efficiently recognize the infected lung images [6] [7] [8] [9]. In the AI-based methods, machine learning and deep learning architectures stands out in most of the COVID-19 classification tasks [10] [11] [12]. But one of the biggest hindrances the researchers are facing is a deficiency in the dataset. To efficiently train a model, a reasonable amount of the subject images is required. But there is an insufficient amount of COVID-19 affected lung available for the research. So, image processing or machine learning model hardly can segregate the COVID and non-COVID images. To support this dataset complication, researchers are lying towards deep learning architecture because of the augmentation and transfer learning approaches [13] [14]. Various versions of CNN models such as — Inception Net, XceptionNet, ResNet, VGGNet, etc. are the prominent architectures that are employed in this research.

Studying the existing classification models, we have adopted High-Resolution Network (HRNet) for feature extraction. In HRNet, different types of convolutional resolutions are linked in a parallel manner. Besides, HRNet consists of plentiful interaction across low and high resolution that bolsters its internal representation ability. The feature representation of this network is kept up during the training process that can prevent the small target information loss in the feature map. For vision-based problems, like small target segmentation and classification, HRNet gives more accurate and definite results because of the parallel procedure. In summary, our contributions in this research are as follows:

– Firstly, we propose a COVID-19 classification method based on a high-resolution network for feature extraction, provides competing results compared with the existing architectures.
– Secondly, we integrate UNet for lung segmentation along with the HRNet to precisely and accurately classify the COVID region. This addition improves the result significantly and validates the infected lung region rather than the redundant non-relevant areas.
– Finally, we conduct a performance comparison with the existing advanced models by implementing those models and evaluating their performance with our proposed work. From the experimental results, we can affirm that the proposed model accomplishes surpassed the existing models by accuracy, sensitivity, specificity, and other evaluation metrics.

The rest of this paper is organized as follows. In Section II, we provided a study on the related works in this area of research. Subsequently, in Section III we discussed our proposed model which consists of the detailed description of our dataset and classification network. In Section IV, the experimental analysis is presented in detail, followed by the performance evaluation. Finally, in Section V, we concluded this paper with significant future works.

## 2 Related Works

Over the years, numerous types of works have been established to detect COVID-19 disease from a distinct perspective. Researchers around the world tried to come up with a model that can efficiently classify this disease considering a short amount of time. In this section, a study on the existing works on COVID-19 classification will thoroughly describe with appropriate characterization and depiction.

Apostolopoulos et al. [14] proposed an architecture based on transfer learning for the feature extraction. Firstly, the authors tried to employed a CNN model to extract the feature of a different nature that is called pre-trained (only used for feature extractor) the CNN model. This was done by operating the transfer learning for feature extraction. After that, the extracted features were fed into a particular network for classification purposes. Though the author accomplished exceptional result but the author did not focus on handling the negative transfer.

Borghesi et al. [15] introduced a chest X-Ray (CXR) scoring system named as Brixia score to determine the outcome (recovery or death). Dividing the lungs into six zones with the aid of frontal chest projection, the authors assigned four scores (Score 0: no abnormalities to Score 3: alveolar predominance or critical condition). Then the six scores for the six divided zones were aggregated to obtain the final score ranging from 0 to 18. For validation, weight kappa *(k*_w_), confidence interval (CI) and P-values were calculated. Although the scoring system is a unique way to identify the disease, the experiment should apply to a considerable amount of CXR images.

Leveraging the multi-resolution capabilities of the inception module, Das et al. [16] built a truncated inception net architecture associating with the adaptive learning rate protocol. In this model, kernels of disparate receptive fields were executed in a parallel manner for feature extraction. Then, the extracted features were deformed depth-wise to obtain the final output. Because of the diminutive dataset, used in the architecture, the inception net is truncated at some particular point of the model. An accuracy of 99.92% was achieved classifying COVID-19 cases combining with the pneumonia cases. A COVID-19 detection model considering multi-class and hierarchical classification was developed by Pereira et al. [17]. Working on the natural data imbalance, a resampling algorithm for rebalancing the class distribution was operated. For feature extraction, Inception V3, Local Binary Pattern (LBP), local phase quantisation (LPQ), Local directional number pattern descriptor (LDN), Locally Encoded Transform Feature Histogram (LETRIST), Binarized Statistical Image Features (BSIF), etc. model was employed. Early and late fusion techniques were leveraged to the feature descriptor algorithms. Then, the author introduced the resampling operation to rebalance the distribution of the multi-class: flat and hierarchical. A macro-average *F_1_* score of 0.65 and 0.89 was achieved using the multi-class approach and the hierarchical classification respectively. In an unbalanced environment, this architecture stands out in the relative existing works.

A 19-layer CNN architecture was proposed by Ozturk et al. [18] for the binary COVID-19 classification. The authors also covered the multiclass problem by assimilating the Pneumonia with the COVID and No Finding class. DarkNet-19, adopted by the author, is based on the prominent real-time object detection model YOLO. In the proposed model, a total of 17 layers were built by a 2D Convolutional layer of different dimensions and trainable parameters, one Flatten and one Linear layer. Each of the Conv. Layer was developed followed by batch normalization and LeakyReLU operation. An accuracy of 87.02% for multi-class cases and 98.08% for binary classes was attained by the authors. A highly diverse and long-rang selective connective method was proposed by Wáng et al. [19]. The machine-driven design strategy is leveraged by generative synthesis [20]. A PEPX module (conv1 × 1 + conv1 × 1 + DWconv3 × 3 + conv1 × 1 + conv1 × 1) was assembled with general convolutional, flatten and fully connected layers. Finally, softmax was used for classification purposes. Though an accuracy of 93.3% was achieved operating this network, the long-range connections in the densely-connected DNN produce memory overhead. Also, the architecture is computationally expensive for the long-range connections in the network. Moreover, heterogeneous incorporation of convolutional layers with different kernels and grouping configurations, heavily affect the interconnection and operation of the architecture.

Narin et al. [21] worked with the pre-trained models such as — ResNet50, InceptionV3 and Inception-ResNetV2. Because of the diminutive amount of data, transfer learning was incorporated to overcome the training time and deficiency in the dataset. Firstly, the input images were fed into the pre-trained models integrated with the transfer learning. Secondly, in the training phase, Global Average Pooling, Fully Connected Layer with ReLU was employed. Finally, the authors concluded with a fully connected layer with softmax for the final classification. The model achieved 97%, 98% and 87% accuracy respectively by operating InceptionV3, ResNet50 and Inception-ResNetV2 architecture. Nevertheless, transfer learning was incorporated with the model for the deficiency in the dataset, the model overfits with the data.

From the studied research works, we have summarized the methods, datasets and performance in Table 1. Though these deep learning methods worked well on this classification, there is a high chance of biasness and oversampling in the learning process for an insufficient number of images. Also, the feature information needs to incorporate in each and every layer on high-to-low and low-to-high upsampling processes which is not integrated with the existing model. To overcome these difficulties, High-Resolution Network (HRNet) is employed for feature extraction of this classification. Moreover, in the existing models, researchers didn’t focus on the segmentation as it plays a critical role to train an architecture. We should exclude the redundant area of a lung image to efficiently work on the infected lung portion only. For that purpose, we have integrated UNet architecture for segmentation purposes. In summary, we have built a unique and unprecedented COVID-19 detection architecture based on UNet (segmentation purpose) and HRNet (feature extraction purpose).

**Table 1:** Analysis of the existing COVID-19 classification works

| Method | Image and Dataset | Performance |
| --- | --- | --- |
| Transfer Learning [14] | 1. 1427 (COVID-19: 224, Pneumonia: 700, Normal: 504)<br>2. 1442 (COVID-19: 224, Pneumonia: 714, Normal: 504) [22], (RSNA), Radiopaedia, and (SIRM) | 1. Accuracy: 96.78%<br>2. Sensitivity: 98.66%<br>3. Specificity: 96.46% |
| Brixia Scoring System [15] | COVID-19: 100 (Departmental Archive) | $k_w$: 0.82; CI: 95% |
| Truncated Inception Net [16] | 1. COVID-19: 162<br>2. Pneumonia 4280 + 1583<br>3. TB (China) 342 + 340<br>and TB (USA) 58 80 [22] [23] [24] | Accuracy of 99.96% (AUC of 1.0) |
| Flat and Hierarchical classification [17] | 1144 CXR images [22] [25] | F1-Score of 0.89 |
| DarkNet + YOLO [18] | COVID-19: 127, Pneumonia: 500, Normal: 500 [22] [25] | Accuracy: 98.08% and 87.02% |
| COVID-Net [19] | 13,975 CXR images [22] [26] [27] | Accuracy: 93.3% |
| ResNet and InceptionNet [21] | COVID-19: 50, Normal: 50 [22] | Accuracy: 97%, 87% |
| VGG19, DenseNet [28] | 50 CXR images with 25 COVID-19 cases [22] | Accuracy: 90%, 90.01% |
| Anomaly Detection [29] | 1431 images (COVID-19: 100) [30] | Accuracy: 96.00% COVID-19 and 70.65% non-COVID-19 |
| DeTraC CNN [31] | 80 Normal, 105 and 11 samples of COVID-19 and SARS [22], JSRT | Accuracy: 95.12% |
| Modified AlexNet [32] | 85 X-ray and 203 CT (BSTI) | Accuracy: 98% and 94.1% (pre-trained) |
| Capsule Network [33] | ImageNet dataset for pre-training | Accuracy: 98.3% and 95.7% (pre-trained) |

## 3 Proposed Model

In this section, we briefly discussed our approaches to data preprocessing, proposed research methodologies with proper illustration and characterization. To build this model, we have explored several preprocessing and classification techniques including supervised machine learning, deep learning, and transfer learning. In the extant research on COVID-19 detection, supervised learning focuses on the binary classification (COVID vs Non-COVID) or a multiclass classification (COVID vs Pneumonia vs Normal lung conditions) [34]. Studying these existing works and considering the drawbacks described in the literature review, we have introduced HRNet [35] for feature extraction embedded with UNet [36] for segmentation purposes. Firstly, this segmentation process has been introduced because COVID chest X-ray images that are publicly available contain several redundant marks, lung regions cropped, shifted to different directions, etc. After that, for feature selection, HRNet has been introduced in the field of COVID-19 detection from CXR images. HRNet has the capability of avoiding small features from the images. After feature selection, a classification head, created of a fully connected neural network has been used to classify COVID vs Non-COVID images. Furthermore, a standard dataset is developed for COVID detection from chest X-ray images from several public data sources. These data sources are updated every day and giving researchers opportunities to focus more on COVID detection from chest X-ray images. A depiction of the proposed architecture is illustrated in Fig. 1. Following, we thoroughly described the related steps of the proposed method.

**Fig. 1:**
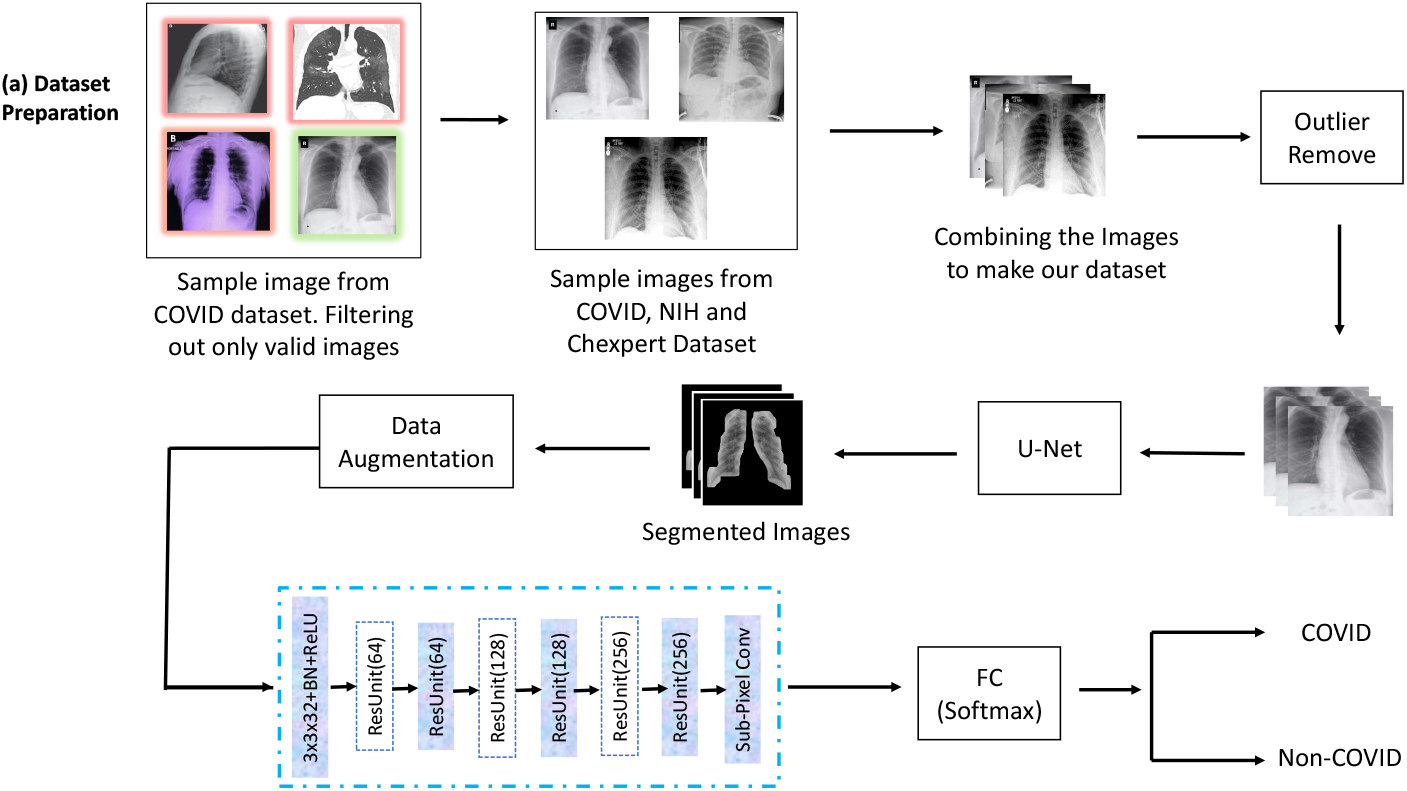
Proposed model to classify COVID-19 disease using HRNet

### 3.1 Dataset Collection and Preperation

The dataset for the classification purpose is built by assembling images from several acknowledgeable sources. Firstly, the COVID-19 Chest X-Ray (CXR) dataset that has been used in our experiment is collected from the public repository of GitHub [22]. As of July 03, 2020, this repository contains 759 images. In Fig. 2, the class distribution of the public repository is depicted. In this public repository, 521 images are labeled as COVID-19 and 12 images are labeled as Acute Respiratory Distress Syndrome (ARDS) COVID-19. A total number of 533 images from the repository were collected from this source for our primary COVID-19 dataset. Though this is the most prominent repository since the beginning of the research in COVID-19 classification, there are some shortcomings in the images. One of the particular drawbacks can be exemplified as — this collection contains images from several publicly available resources such as websites and pdf formats. As a result, these images are in variable size and quality. Also, there are few side view images where the majority of the images belong to frontal views. Moreover, some images have color issues, contain markers, etc. In Fig. 3, four examples of COVID-19 images are depicted acknowledging the issues — side view (Fig. 3a), washed out (Fig. 3b), color issue (Fig. 3c) and markers (Fig. 3d).

**Fig. 2:**
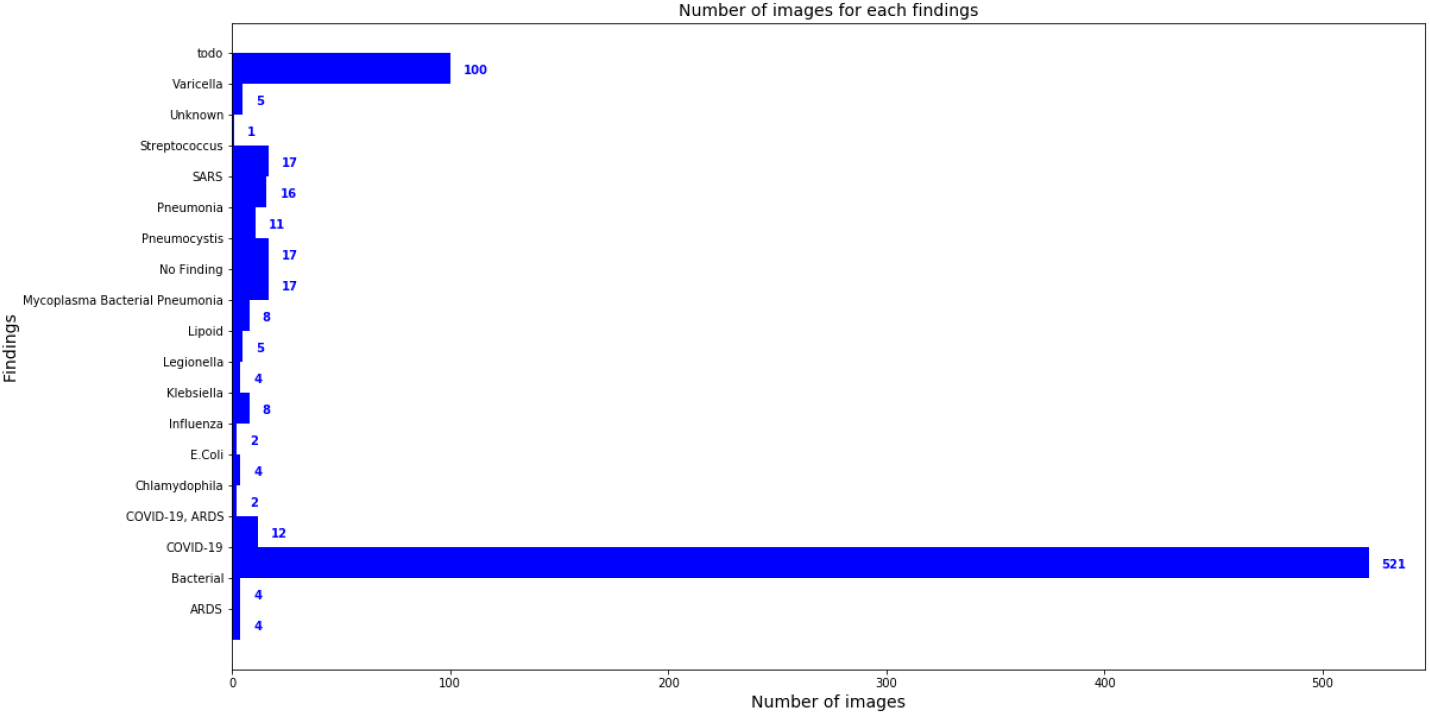
A class distribution representation of the public repository.

**Fig. 3:**
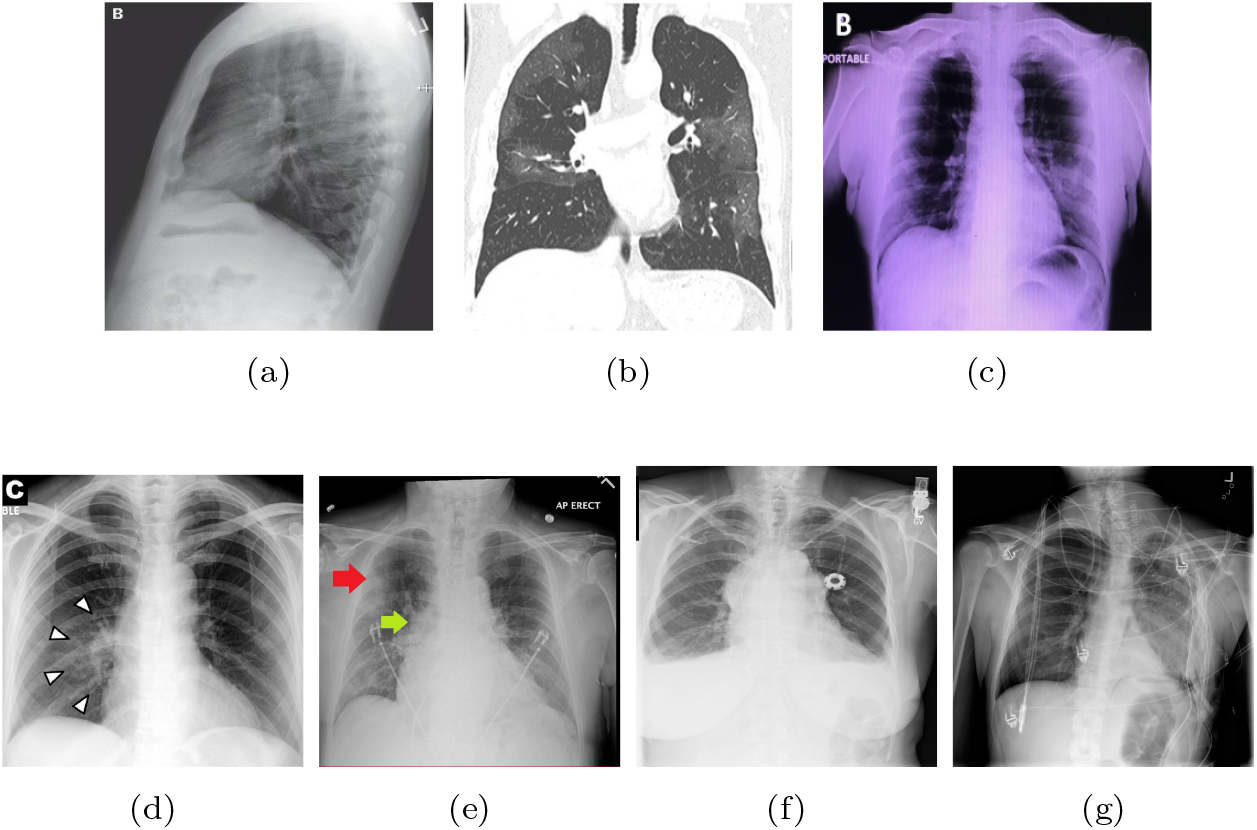
Variations observed in COVID dataset [(a) side view, (b) washed out, (c) color issue, (d) and (e) markers, (f) and (g) medical equipment]

For non-COVID/normal images, we have collected images from the National Institute of Health (NIH) Chest X-Ray [25] which contains 108,948 frontal view X-ray images with 14 condition labels including normal condition and pneumonia of 32,717 unique patients. Unlike the COVID dataset, these images are not of variable dimensions. All of the images are resized to 1024 x 1024 in portrait orientation. As most of the images from the COVID dataset belongs to adult patients, we have applied an age threshold of 18 years or older on normal condition images to keep the X-Ray image condition as similar as possible. We also explored the ChexPert [37] which contains 224,316 chest radiographs. This dataset contains 14 labeled observations of 65,240 patients where the number of Normal (non-COVID) images are 16,627.

As described earlier, COVID images contain lateral X-ray images, taken from PDF files, marked images (fig. 3d and 3e), etc. Firstly, we removed these images from the COVID dataset as there is a possibility that these images can make the classifier biased. Secondly, in NIH and ChexPert datasets, some images contain medical equipment (fig. 3f and 3g) which creates redundancies and abnormalities in times of training the model. Hence, these types of images are excluded from the main data repository. But still, most of the images contain several marks around the lungs area. To avoid these marks, we segmented the lung area from the images. In the next section, we described the process of lung segmentation and preprocessing the images and finally creating a practical COVID dataset for our experiment. In fig. 4, a comparison between a single random image collected from COVID, NIH, and ChexPert dataset is illustrated. Furthermore, we aggregated Normal images from NIH and ChexPert dataset as it looks similar. In Table 2, we have summarized the properties of the final dataset created.

**Fig. 4:**
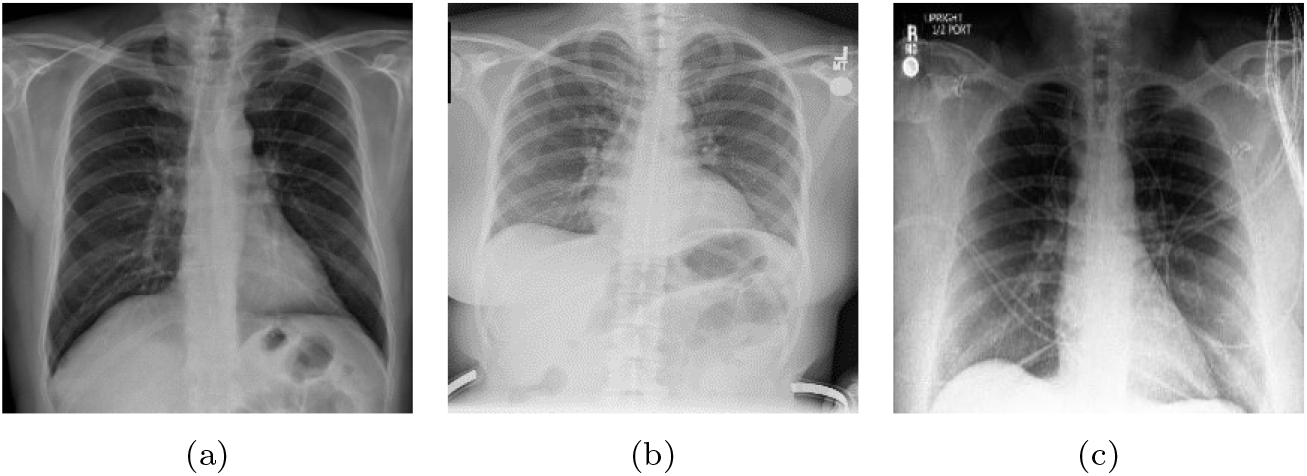
Image comparison of COVID and non-COVID dataset [(a) COVID dataset (b) NIH dataset, and (c) ChexPert dataset]

**Table 2:** Primary data selection

| Dataset Name | Number of Images | Image Characteristics | Selection Criteria |
| --- | --- | --- | --- |
| Covid Public Repository | 472 | Variable size, brightness, contrast | All COVID images. |
| NIH Chest X-Ray | 1000 | Constant size of $1024 \times 1024$, Uniform | No findings, age threshold, random selection, without any medical equipment |
| ChexPert | 1000 | Variable size, resized to $256 \times 256$ | No findings, age threshold $> 18$, random selection, without any medical equipment |

### 3.2 Lung Segmentation

For our primary purpose, all the datasets mentioned above do not contain any annotation for the lung area. Thus to segment the lung data, we collect dataset that has lung area annotated and train the UNet [36] from scratch. Belongs to the semantic segmentation category, UNet was solely created for medical image segmentation and also proven its worth in recent segmentation tasks. Another great feature of UNet is it can make a strong usage of augmented data, contracting and symmetric path based architecture enables precise localization.

Assembled upon the Fully Connected Network (FCN), UNet is symmetric and consists of a skip connection which provides information from local to global network while upsampling. As a consequence of the symmetricity, this network has an extended number of feature maps in the intermediate connection that allows transferring the information. There are three distinct parts of the UNet — Downsampling path (The Contraction), Bottleneck, and Upsampling path (The Expansion). While the Contracting path is established by the basic convolutional process, the Extracting path is constituted with transposed 2D convolutional layers.

For this segmentation task, we collected a dataset from Jaeger et al. [38]. This dataset contains a total number of 800 frontal X-ray images from the Montgomery County chest X-ray dataset of the Department of Health and Human Services, Montgomery County, Maryland, USA (138 images) and Shenzen chest X-ray dataset of Guangdong Medical College, Shenzen, China (662 images). Chest X-ray datasets from these sources are visually similar to the dataset of 2472 images which we have selected for our COVID detection task. A side by side comparison with our COVID dataset images and segmentation dataset images is depicted in Fig. 5.

**Fig. 5:**
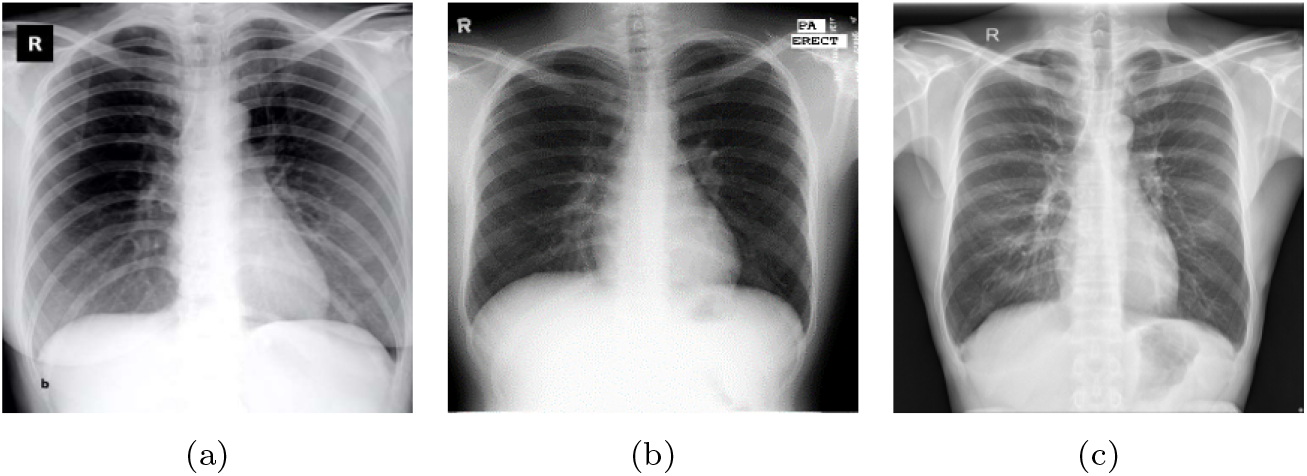
Image comparison used in lung segmentation [(a) COVID dataset, (b) Montgomery dataset, and (c) Shenzen dataset]

Every image of the Montgomery dataset is either 4020 × 4892 pixels or 4892 × 4020 pixels. Images from the Shenzen dataset are variable but approximately 3000 × 3000 pixels. We resized these images and their corresponding lung masks to 512 × 512 pixels to train our model. Moreover, augmentation is done on the images and their corresponding masks to take the advantages of the UNet model. The augmentation procedure and parameters are described in the dataset augmentation section and the training process with the parameters is explained in the Experimental Analysis section. After training the UNet model with this segmentation dataset, we used the operating weights to get segmented lung images from the COVID dataset. Furthermore, the UNet model provides a prediction mask for every image. Hence, we applied dilation operation for 8 iterations and 2 iterations of closing operation on the predicted masks to gain more information from the edges of the lungs. To ensure the quality of the segmentation, we then applied a region-based threshold to distinguish the pixels. For every image, we got several segmented regions then calculate the total area of these segmented regions for image and apply a threshold of 50,000 pixels to ensure the quality. This threshold value was chosen by trial and error and observing the output from the segmentation model. After applying an area threshold to these segmentations, we finally got our COVID dataset. Our final COVID dataset consists of 41G COVID images and 5GG non-COVID images from NIH and the ChexPert dataset. The working procedure of this segmentation method is summarized in Algorithm 1.

**Fig. 6:**
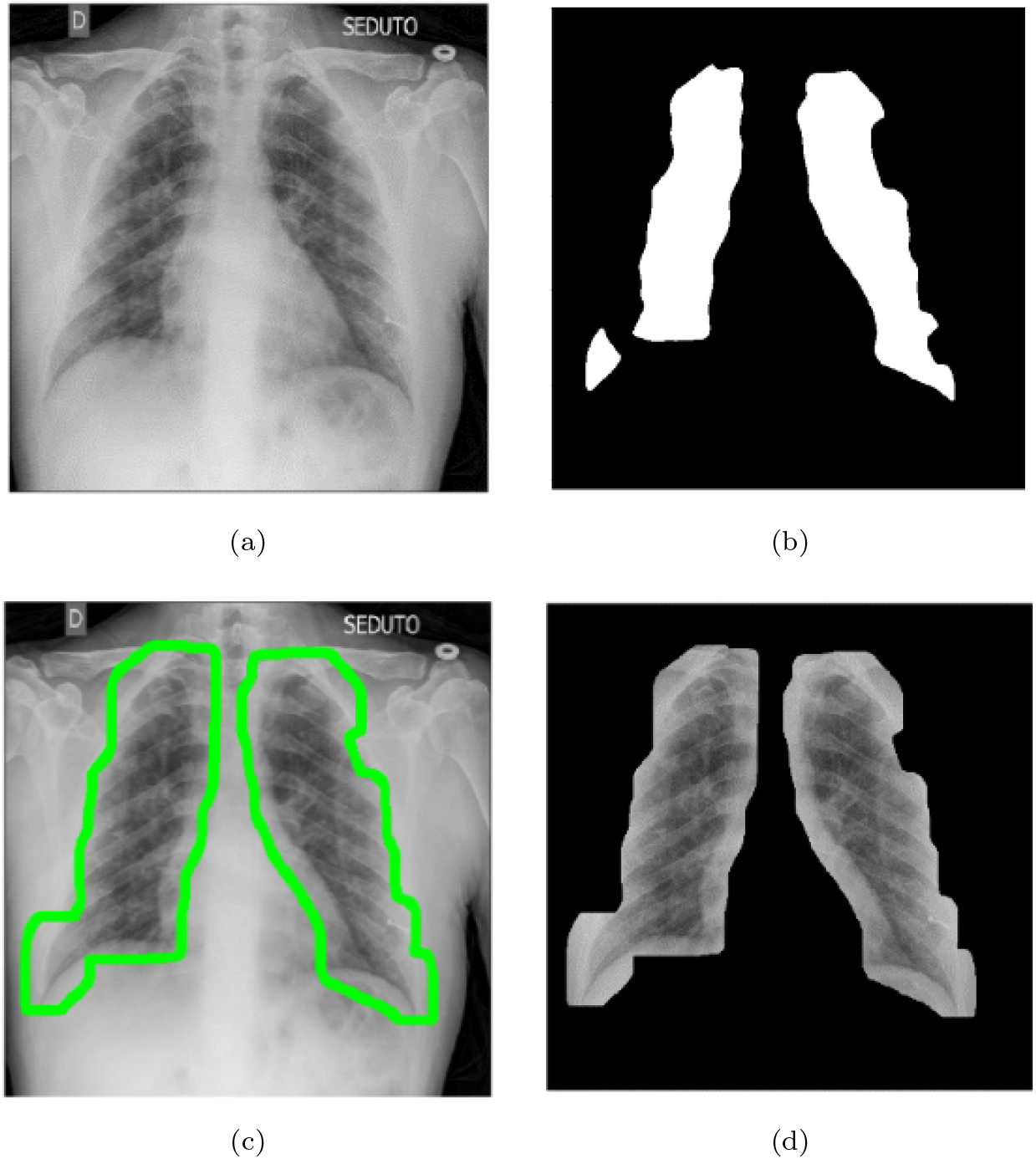
Segmented region after dilation and closing apply UNet architecture

### 3.3 Data Augmentation

For a better generalization of the classification model, we augmented both the segmentation dataset for the UNet model and the preprocessed segmented COVID dataset. In both cases, we applied the same set of augmentation such as scaling, padding, crop, rotation, gamma correction, slight Gaussian blur, random noise, salt and pepper noise, etc. based on a probabilistic value. Fig. 7 demonstrates the output of image augmentation. In Table 3, we have discussed the parameters of augmentation. These parameters were used for both the segmentation dataset and the COVID dataset. All of these parameters were selected empirically.

**Algorithm 1:**
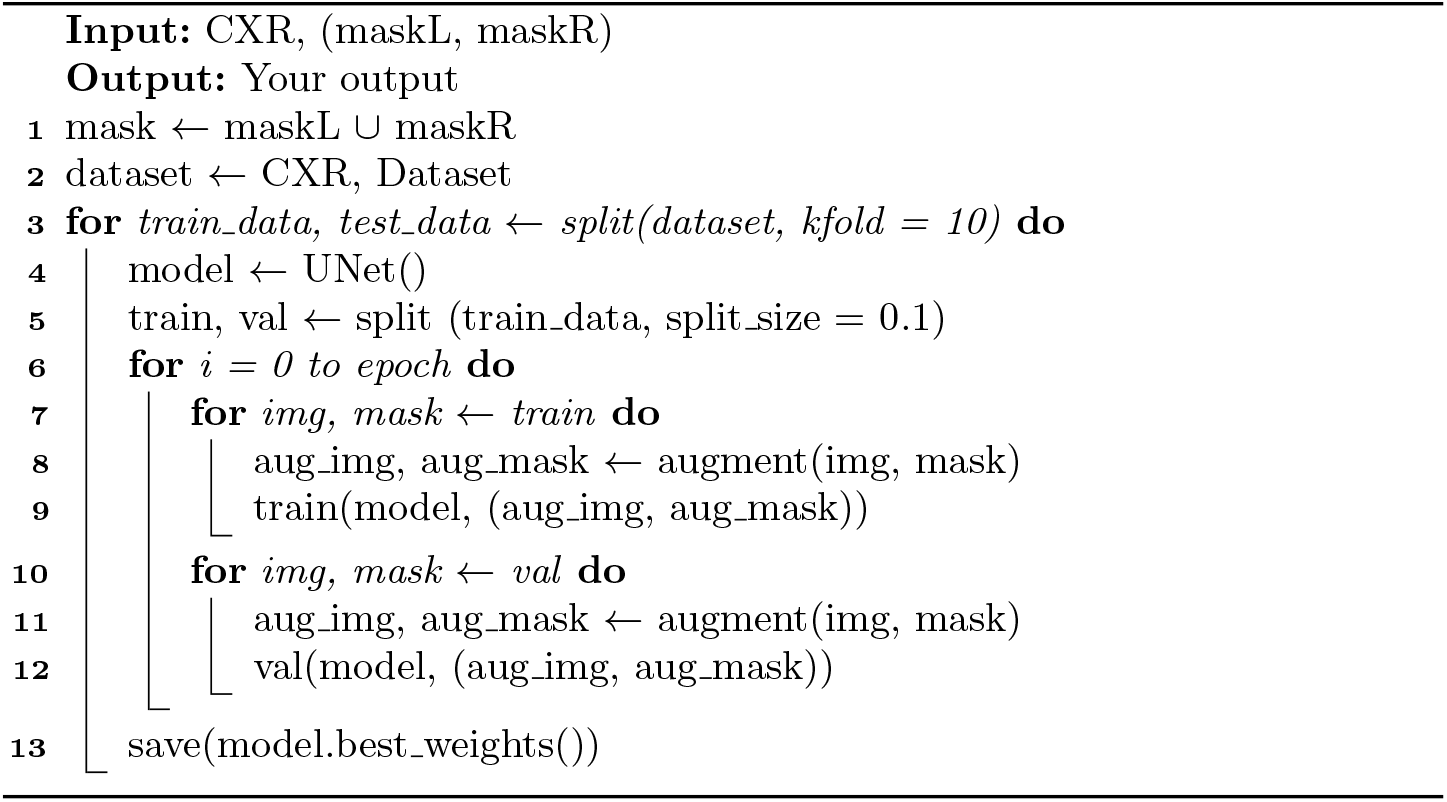
Lung Segmentation

**Fig. 7:**
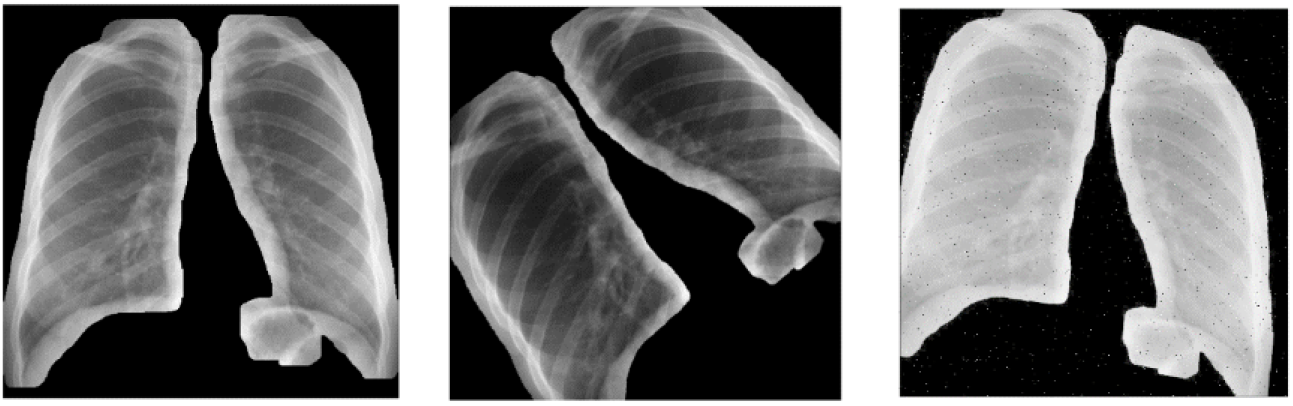
Sample images after augmentation.

**Table 3:** Image augmentation parameters

| Process | Values | Probability |
| --- | --- | --- |
| Scale | X-axis – (0.8 to 1.1)<br>Y-axis – None | 1 |
| Rotate | Angle – (-45, 45) | 1 |
| Shear | X-axis – (-0.2, 0.2)<br>Y-axis – None | 0.7 |
| Padding | 512 | 1 |
| Center Crop | 512 | 1 |
| Gamma Correction | 0.5 to 3 | 1 |
| Salt and Pepper Noise | 0.01 | 0.3 |
| Blur | 0.5<br>Kernel size = (11, 11) | 0.5 |
| Random Noise | 0.5 | 0.3 |

### 3.4 Classification with HRNet

For feature extraction, High Resolution Net (HRNet) [35] is a state-of-the-art neural network architecture. In most of the recent HRNet adopted architecture, it is used as the backbone of the proposed models. Considering two approaches—Top-Down and Bottom-Up, HRNet followed the top-down approach because it detects the subject first, establishes a bounding box around the subject, and then estimate the significant feature. Moreover, HRNet relies on continuous multi-scale fusions instead of a single high-to-low upsampling process. In the following, a brief description of the HRNet architecture is characterized.

The fundamental working procedure is it calculates lower resolution and higher resolution sub-network parallelly. Then, these two networks coalesced together by a fuse layer for the purpose to assemble and interchange information from each of the sub-network with each other. Consisting of four parts, each of the parts is built with repeating modularized multi-resolution sections [35]. Each section consists of a group convolution supporting the multi-resolution properties. A multi-resolution convolution can be constructed with the aid of regular convolution wherein a regular convolution, the input, and output layers are connected in a fully-connected approach [39]. The subnetworks follow these properties to aggregate their multi-resolution attributes. In this way, the subnetwork gains global high-resolution representations. A representation of the employed HRNet architecture is depicted in Fig. 8. The working procedure of feature extraction using HRNet and classification is compiled in Algorithm 2.

**Fig. 8:**
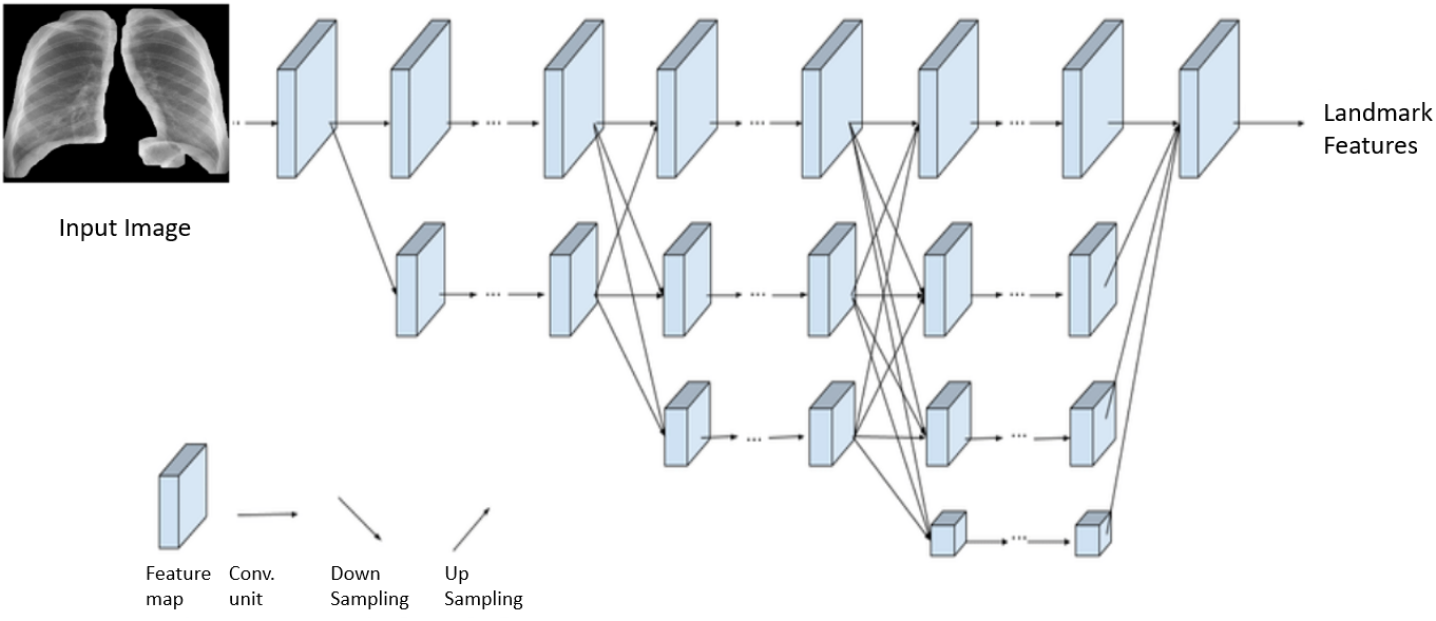
A general architecture of HRNet for feature extraction

**Algorithm 2:**
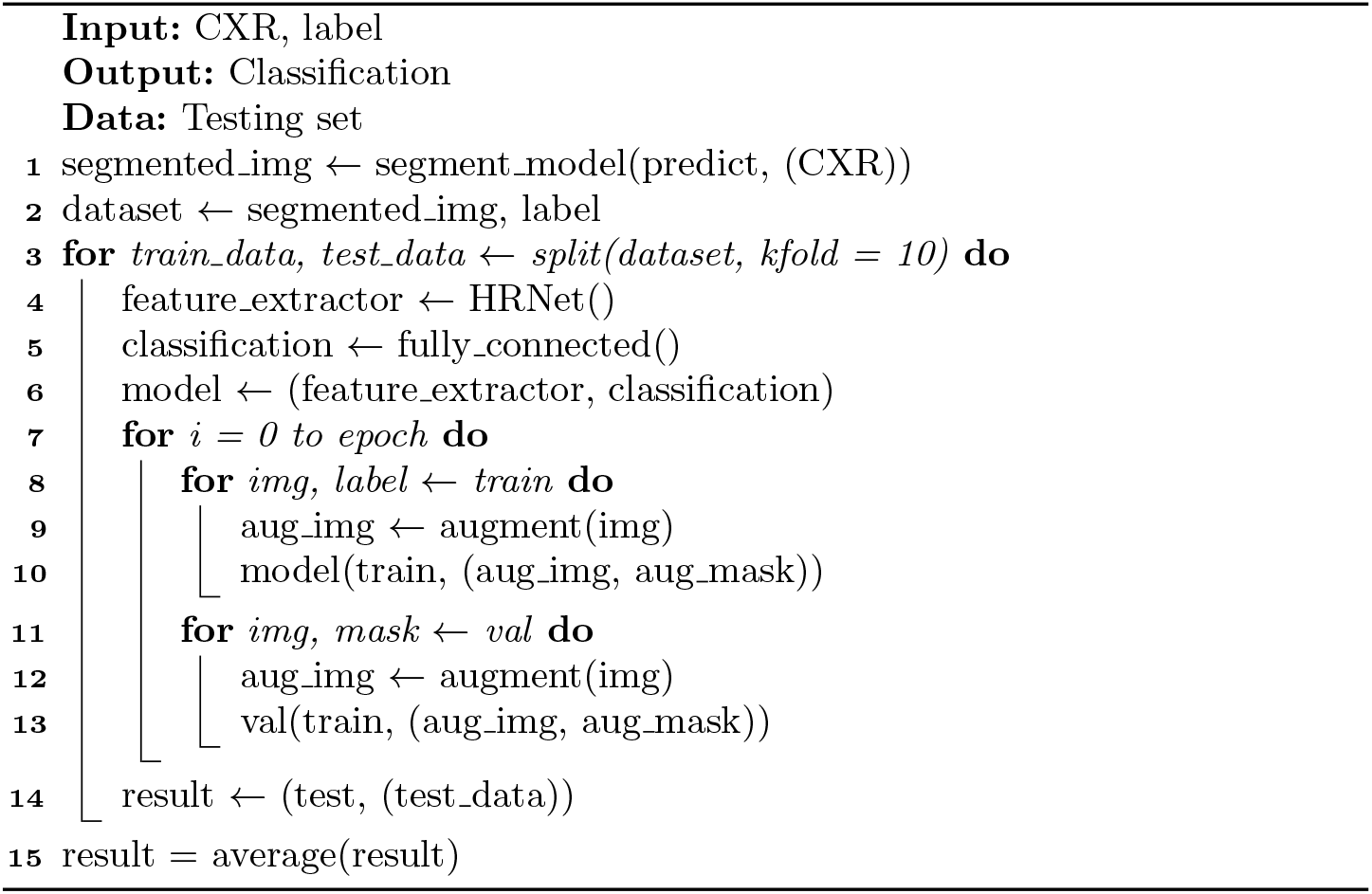
COVID-19 Classification

## 4 Experimental Analysis

### 4.1 Computing Infrastructure

This whole experiment was conducted in a device with Ryzen 7 3700X processor, NVIDIA GeForce RTX 2070 Super GPU, 16 GB 3600 MHz RAM, and Samsung Evo 970 m.2 SSD.

### 4.2 Dataset Segmentation

To make an accurate detection, the classifiers need to focus on the lung regions properly. Our previous experiments without segmentation. Fig. 9 shows that despite having high accuracy on training and validation set, the focus region of the classifiers often deviate to outside of lungs which may lead to a false prediction. To address this issue we segmented our dataset and kept only the lung portion from the X-ray images.

**Fig. 9:**
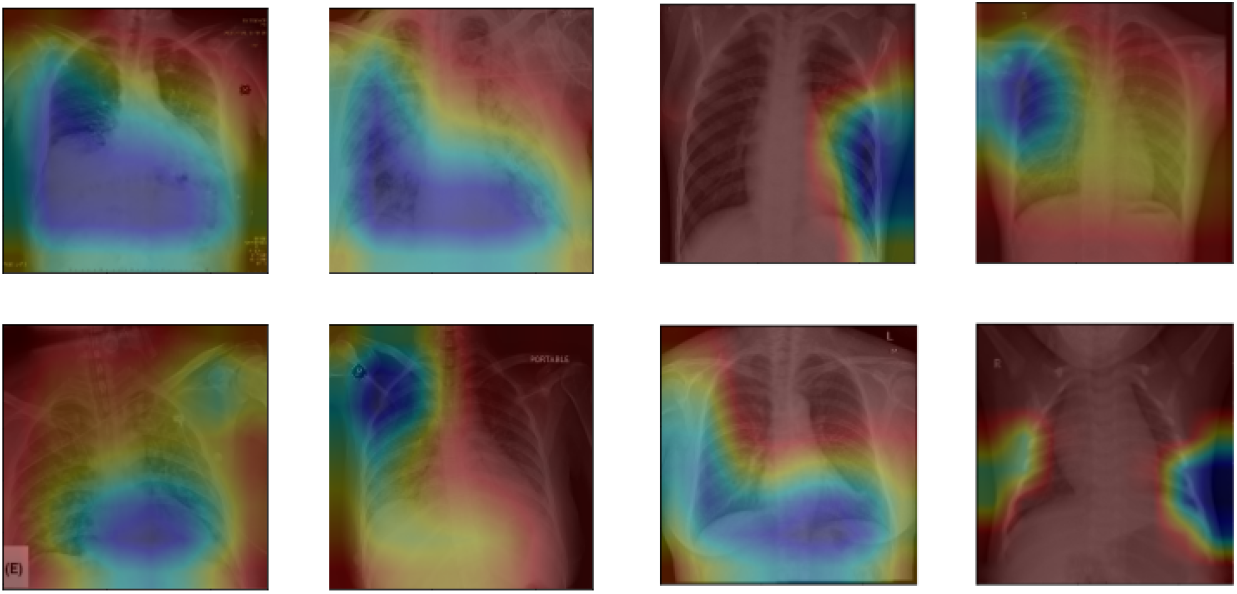
This figure visualizes some sample heatmaps (blue shade represents the focus region) of corona positive detection by the classifier. But as we can see in the figure, most of the focused regions redundant, not significant, erroneous and somewhat can produce a biased result in times of classification.

Only lung areas are collected to create a clean dataset without any redundant markers and flaws in the image. To collect these lung areas from the primary dataset of 2472 images, the UNet model was trained on Montgomery and Shenzen dataset. This dataset has 800 images and to train UNet KFold (10 Fold) crossvalidation is used. In this dataset, two corresponding masks, one for the right lung and one for the left lung contains for one CXR image. At first, two lung images were combined to make one corresponding mask for each CXR image. These images are then employed to train the UNet model. For the loss function, a Dice coefficient loss is used to get crisp borders. This loss function has been introduced by Milletari et al. [40] in their research of 3D volumetric image segmentation. Dice loss originates from Sørensen-Dice coefficient. It is a statistical method developed in 1940. Given dice coefficient D, it can be written as,

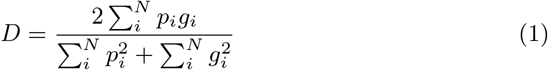

Here *P_i_* and *G_i_* represents the pixels of prediction and ground truth respectively. In edge detection, the values of *P_i_* or *G_i_* is either 0 or 1. That means if two sets of pixels overlap perfectly the *D* reaches its maximum value to 1, otherwise it decreases. By using dice loss, two sets of predicted edge and ground truth edge are trained to overlap gradually. After that, traditional cross-entropy loss calculates the average of per-pixel loss where the per-pixel loss is calculated discretely here without knowing if the adjacent pixels are edges or not. As a result, it is not enough for image-level prediction. Dice loss provided better results in our lung segmentation using UNet. The model was trained for 25 epochs for each fold with a learning rate of 0.005, and a custom dice coefficient loss function with a grayscale input image of size 512 × 512 pixels. Table 4 shows the input, output, and layer configuration used for UNet.

**Table 4:** UNet Configuration

| Layer | Parameters | Activation | Output Shape |
| --- | --- | --- | --- |
| Input | — | — | $512 \times 512 \times 1$ |
| Conv2d_1a | $512 \times 512 \times 1$ | ReLU | $512 \times 512 \times 32$ |
| Conv2d_1b | $512 \times 512 \times 32$ | ReLU | $512 \times 512 \times 32$ |
| MaxPooling2d_1 | $512 \times 512 \times 32$ | - | $256 \times 256 \times 32$ |
| Conv2d_2a | $256 \times 256 \times 32$ | ReLU | $256 \times 256 \times 64$ |
| Conv2d_2b | $256 \times 256 \times 64$ | ReLU | $256 \times 256 \times 64$ |
| MaxPooling2d_2 | $256 \times 256 \times 64$ | - | $128 \times 128 \times 64$ |
| Conv2d_3a | $128 \times 128 \times 64$ | ReLU | $128 \times 128 \times 128$ |
| Conv2d_3b | $128 \times 128 \times 128$ | ReLU | $128 \times 128 \times 128$ |
| MaxPooling2d_3 | $128 \times 128 \times 128$ | - | $64 \times 64 \times 128$ |
| Conv2d_4a | $64 \times 64 \times 128$ | ReLU | $64 \times 64 \times 256$ |
| Conv2d_4b | $64 \times 64 \times 256$ | ReLU | $64 \times 64 \times 256$ |
| MaxPooling2d_4 | $64 \times 64 \times 256$ | - | $32 \times 32 \times 256$ |
| Conv2d_5a | $32 \times 32 \times 256$ | ReLU | $32 \times 32 \times 512$ |
| Conv2d_5b | $32 \times 32 \times 512$ | ReLU | $32 \times 32 \times 512$ |
| Conv2D_trans_1 | $32 \times 32 \times 512$ | - | $64 \times 64 \times 256$ |
| Concatenate | Conv2D_trans_1, Conv2d_4b |  |  |
| Conv2d_6a | $64 \times 64 \times 512$ | ReLU | $64 \times 64 \times 256$ |
| Conv2d_6b | $64 \times 64 \times 256$ | ReLU | $64 \times 64 \times 256$ |
| Conv2D_trans_2 | $64 \times 64 \times 256$ | - | $128 \times 128 \times 128$ |
| Concatenate | Conv2D_trans_2, Conv2d_3b |  |  |
| Conv2d_7a | $128 \times 128 \times 256$ | ReLU | $128 \times 128 \times 128$ |
| Conv2d_7b | $128 \times 128 \times 128$ | ReLU | $128 \times 128 \times 128$ |
| Conv2D_trans_3 | $128 \times 128 \times 128$ | - | $256 \times 256 \times 64$ |
| Concatenate | Conv2D_trans_3, Conv2d_2b |  |  |
| Conv2d_8a | $256 \times 256 \times 128$ | ReLU | $256 \times 256 \times 64$ |
| Conv2d_8b | $256 \times 256 \times 64$ | ReLU | $256 \times 256 \times 64$ |
| Conv2D_trans_4 | $256 \times 256 \times 64$ | - | $512 \times 512 \times 32$ |
| Concatenate | Conv2D_trans_4, Conv2d_1b |  |  |
| Conv2d_9a | $512 \times 512 \times 64$ | ReLU | $512 \times 512 \times 32$ |
| Conv2d_9b | $512 \times 512 \times 32$ | ReLU | $512 \times 512 \times 32$ |
| Conv2d_10 | $512 \times 512 \times 32$ | ReLU | $512 \times 512 \times 1$ |

As mentioned above, UNet is trained for 10 Folds, 25 epochs for each fold, and the dice coefficient loss function is used. The learning rate is selected by train and error process and images are augmented according to the configuration described in the augmentation section. Fig. 10 shows the average training accuracy, training loss, average validation accuracy, and average validation loss for 10 folds.

**Fig. 10:**
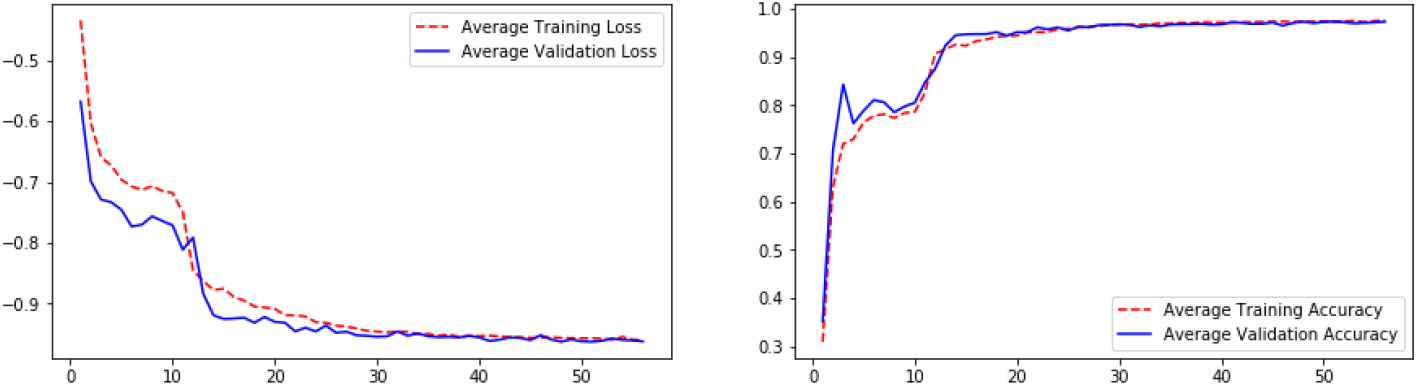
Training and Validation curves after training the UNet architecture

After training the model, the weights are applied on the COVID dataset to remove redundant areas from chest X-ray images and to get lung areas. This segmented dataset is used for training the HRNet and get classification results from the classifier.

### 4.3 Feature Extraction and Training Procedure

In this section, the training procedure of feature extraction using HRNet and classification been discussed briefly. HRNet has the capability of avoiding the loss of small target information in the feature map due to its convolutions being connected in parallel and high-resolution feature representation. To train the model, the segmented COVID dataset of 910 images is used. This dataset contains 410 lungs segmented COVID images and 500 lung segmented non-COVID images. For training purposes, 10 fold cross-validation is used. The model showed better accuracy for learning rate 0.01, weight decay 0.0001, momentum 0.8, learning rate factor 0.1, learning rate step 30, 60, 90. Stochastic gradient descent and binary cross-entropy loss functions are used. These parameters were chosen by observing the performance and evaluating the model progressively (Table 5). The performance analysis of the model is shown in Table 6 for each fold. Using the KFold algorithm, first, the dataset is divided into 10 folds where 1 fold is kept for testing. The other 9 folds are used for training purposes.

**Table 5:**
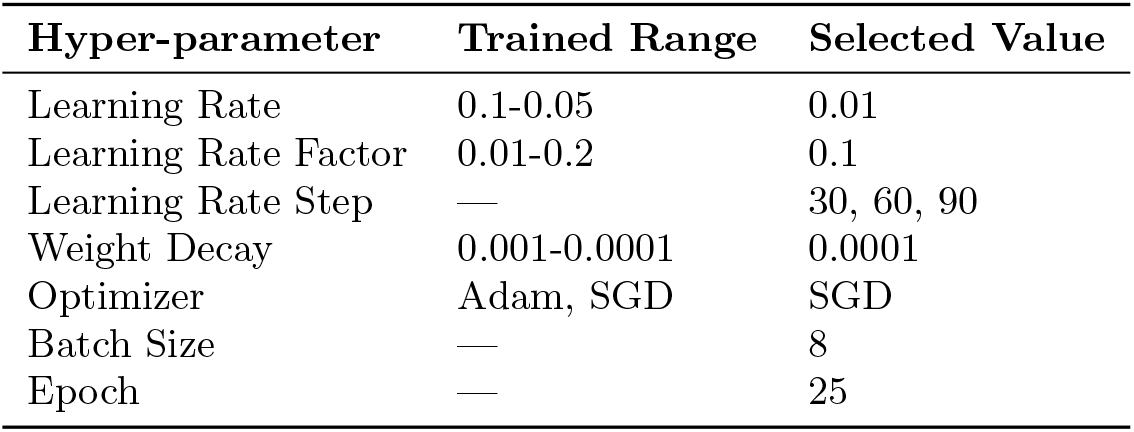
Hyper-parameter Setup

**Table 6:** Performance analysis of the proposed architecture

| Fold Number | Testing Accuracy (%) | Testing Sensitivity (%) | Testing Specificity (%) |
| --- | --- | --- | --- |
| 1 | 98.78 | 97.56 | 98.03 |
| 2 | **100.0** | **100.0** | **100.0** |
| 3 | 98.78 | 97.56 | 98.03 |
| 4 | 98.78 | 97.56 | 98.03 |
| 5 | **100.0** | **100.0** | **100.0** |
| 6 | **100.0** | **100.0** | **100.0** |
| 7 | **100.0** | **100.0** | **100.0** |
| 8 | 98.78 | 97.56 | 98.03 |
| 9 | **100.0** | **100.0** | **100.0** |
| 10 | 97.80 | 95.12 | 96.15 |
| **Average** | **99.29** | **98.53** | **98.82** |

In summary, HRNet has been used as the backbone of our classification model to extract features from images. These features are then passed through several fully connected layers which are defined as a classification head. The parameters of this classification head such as the number of layers, dropout, activation function, regularization, etc. were selected by trial and error method. The configuration of the classification head is given in the fig. 11. Furthermore, for performance evaluation, if the number of True Positive, True Negative, False Positive and False Negative denoted by *TP,TN, FP* and *FN* respectively, then the three adopted evaluation matrices which are as follows:

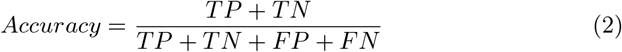

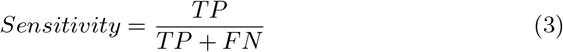

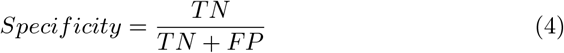

**Fig. 11:**
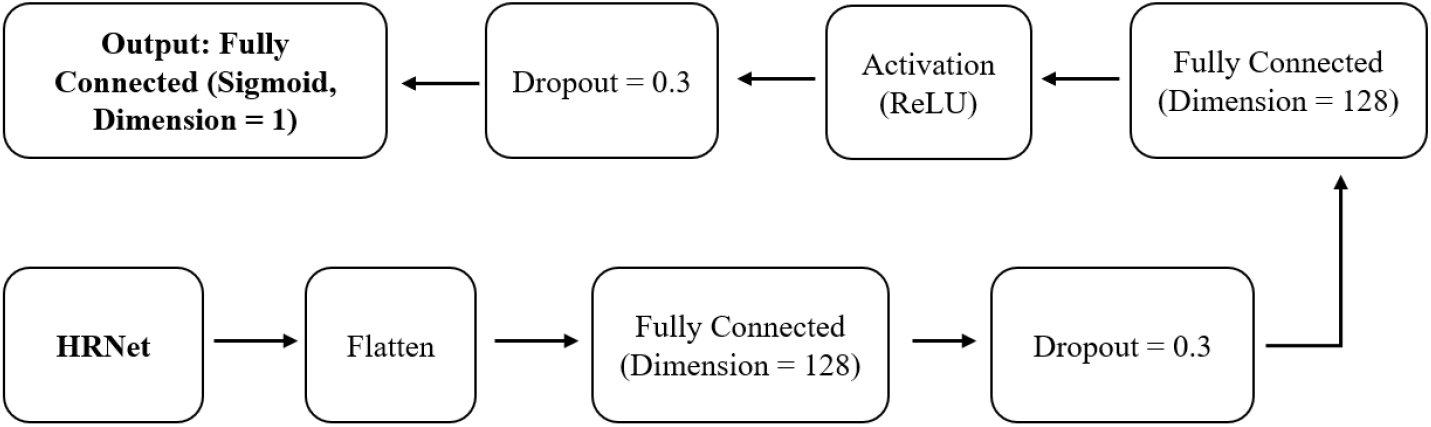
Classification Head

In Table 6, the average of the testing accuracy, sensitivity and specificity for the 10 folds is characterized. We accomplished 99.26% testing accuracy, 98.53% of testing sensitivity and 98.52% of testing specificity. Furthermore, we depicted the confusion matrices of worst and best cases in fig. 13. In the worst confusion matrix(fig. 13a), 39 and 50 testing images correctly classified as COVID and non-COVID respectively whereas only two images are falsely classified. But, in fig. 13b, all the testing images are accurately classified as COVID and non-COVID which represents the best confusion matrix. We have also successfully eradicated the issue of false focused region detection through segmentation. Fig. 12 shows the heatmaps of correctly classified regions.

**Fig. 12:**
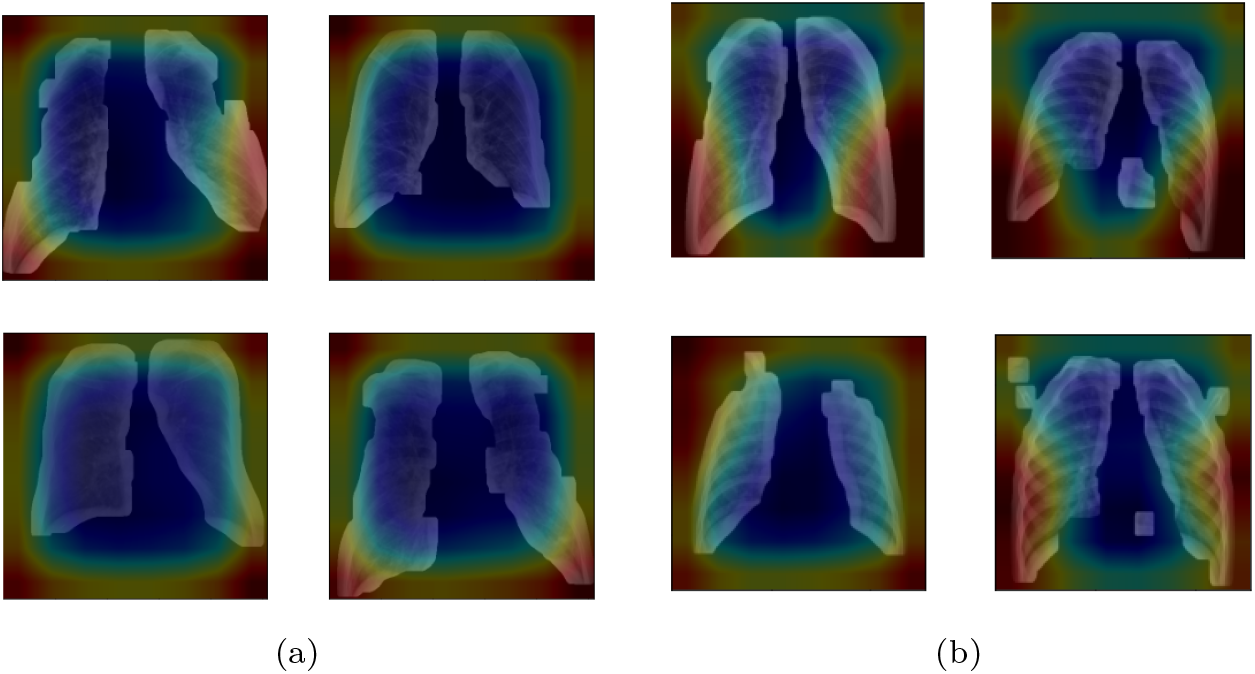
Heatmaps in (a) visualize some sample for positive detection and in (b) represent corona negative detection. As we can see, the lung regions are accurately focused (blue shade represents the focus region) by the classifier after training with segmented data.

**Fig. 13:**
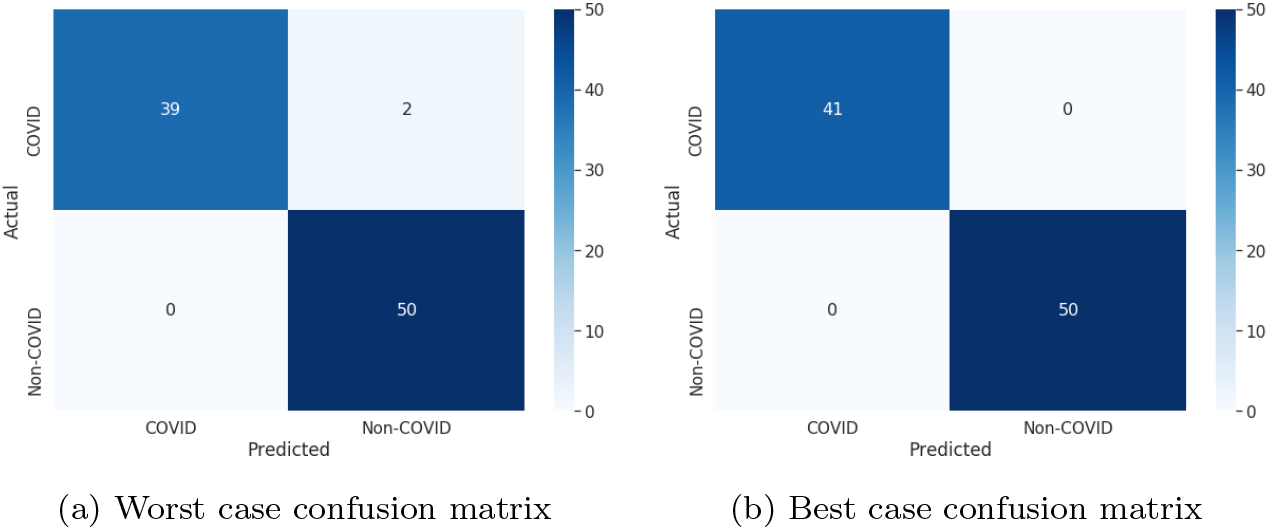
Confusion Matrix after training the proposed model

Moreover, we have compared the performance of the proposed model with the existing adopted models. Three existing prominent architecture — ResNet152, DenseNet121, and EfficientNetB4 are implemented for the comparison purpose. We achieved the superlative result in each of the terms of the evaluation metric by comparing our proposed model with these trained architectures. In Table 7, we have summarized the existing model’s performance comparison with our proposed model. In fig. 14, a depiction of the optimal confusion matrix of the existing trained model is illustrated.

**Table 7:**
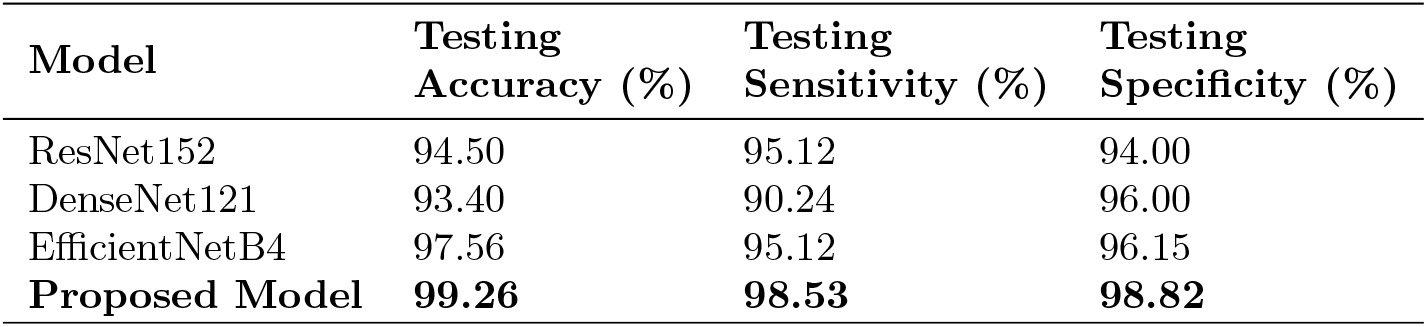
Performance comparison with the existing models

**Fig. 14:**
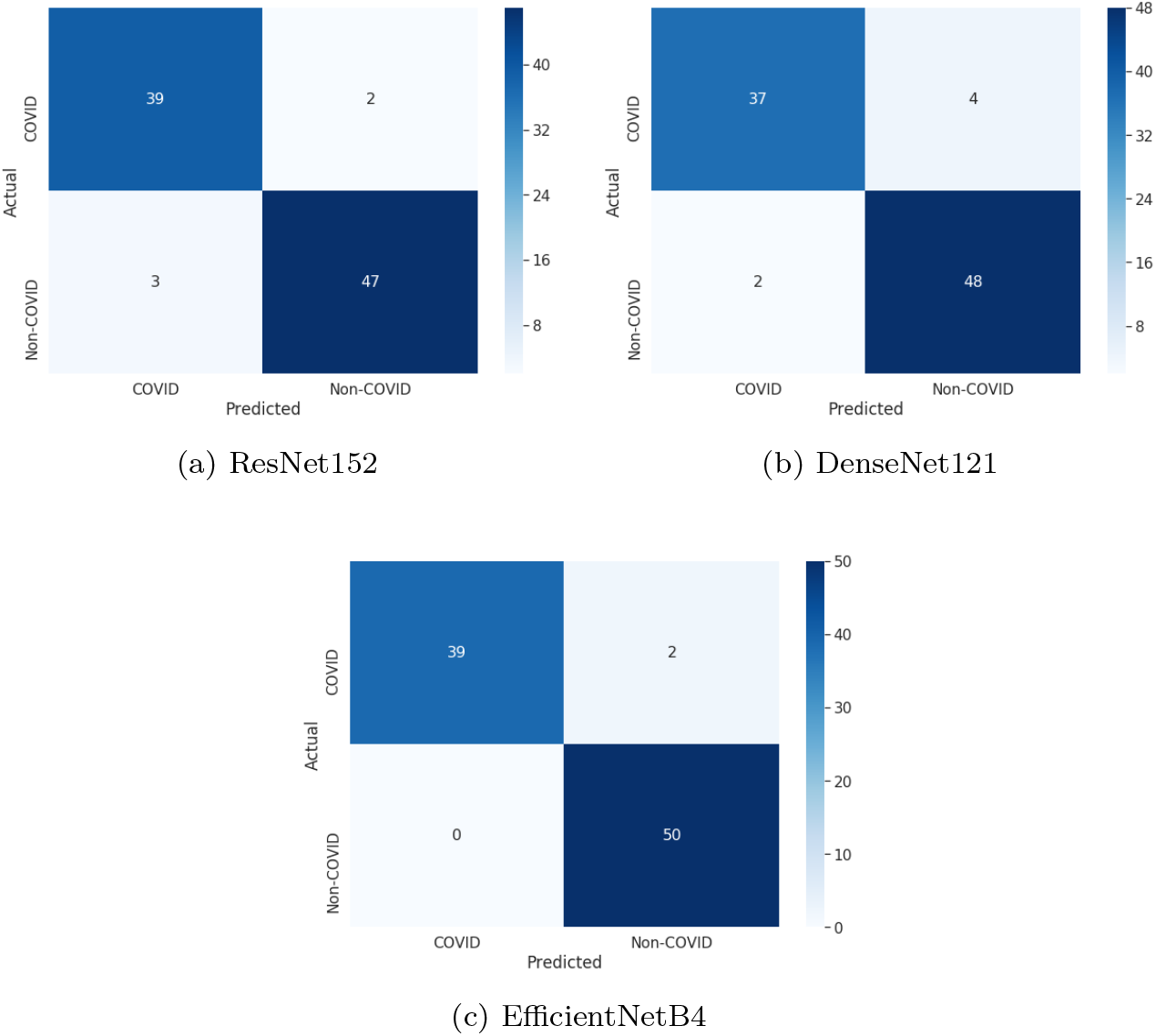
Confusion Matrix after training the existing models

## 5 Conclusion

In this study, we use a segmented X-ray dataset, as extensive experiments show that, with a non-segmented dataset, classifiers may focus outside of the lung region which can lead to false classification results. Meanwhile, we also evaluate some state-of-the-art recognition methods on our dataset. The result demonstrates that HRnet performs the best among the others with 99.26% accuracy, 98.53% sensitivity, 98.82% specificity. The proposed model is fully automated without any need for manual feature extraction. Moreover, to ensure a production-ready solution, we broadly investigate the results and focus regions of the classifiers and our experimental results show the robustness of our proposed model to focus on the right region and classify. To conclude, this model can be used to help the radiologist to make clinical decisions, due to its unbiased high-accuracy and correctly identified focus region. We hope that our proposed methodology is a step towards the possibility of lessening the false positive detection from X-Ray images.

However, in terms of data, we are still in the primary level of the experiment. As the number of patients increasing around the world and the symptoms and formation of the virus are changing day by day, with the continuous collection of data, we intend to extend the experiment further and enhance the usability of the model.

## Data Availability

All the data were collected from several research works which were conducted previously and publicly available data resources,

## References

1. C.-C. Lai, T.-P. Shih, W.-C. Ko, H.-J. Tang, and P.-R. Hsueh, “Severe acute respiratory syndrome coronavirus 2 (sars-cov-2) and corona virus disease-2019 (covid-19): the epidemic and the challenges,” International journal of antimicrobial agents, p. 105924, 2020.

2. Q. Li, X. Guan, P. Wu, X. Wáng, L. Zhou, Y. Tong, R. Ren, K. S. Leung, E. H. Lau, J. Y. Wong et al., “Early transmission dynamics in wuhan, china, of novel coronavirus—infected pneumonia,” New England Journal of Medicine, 2020.

3. C. Huang, Y. Wang, X. Li, L. Ren, J. Zhao, Y. Hu, L. Zhang, G. Fan, J. Xu, X. Gu et al., “Clinical features of patients infected with 2019 novel coronavirus in wuhan, china,” The lancet, vol. 395, no. 10223, pp. 497—506, 2020.

4. D. Chang, M. Lin, L. Wei, L. Xie, G. Zhu, C. S. D. Cruz, and L. Sharma, “Epidemiologic and clinical characteristics of novel coronavirus infections involving 13 patients outside wuhan, china,” Jama, vol. 323, no. 11, pp. 1092—1093, 2020.

5. K. Kuba, Y. Imai, S. Rao, H. Gao, F. Guo, B. Guan, Y. Huan, P. Yang, Y. Zhang, W. Deng et al., “A crucial role of angiotensin converting enzyme 2 (ace2) in sars coronavirus—induced lung injury,” Nature medicine, vol. 11, no. 8, pp. 875—879, 2005.

6. F. Shi, J. Wang, J. Shi, Z. Wu, Q. Wang, Z. Tang, K. He, Y. Shi, and D. Shen, “Review of artificial intelligence techniques in imaging data acquisition, segmentation and diagnosis for covid-19,” IEEE reviews in biomedical engineering, 2020.

7. T. Ebihara, R. Endo, X. Ma, N. Ishiguro, and H. Kikuta, “Detection of human coronavirus nl63 in young children with bronchiolitis,” Journal of medical virology, vol. 75, no. 3, pp. 463–465, 2005.

8. O. Albahri, A. Zaidan, A. Albahri, B. Zaidan, K. Abdulkareem, Z. Al-qaysi, A. Alamoodi, A. Aleesa, M. Chyad, R. Alesa et al., “Systematic review of artificial intelligence techniques in the detection and classification of covid-19 medical images in terms of evaluation and benchmarking: Taxonomy analysis, challenges, future solutions and methodological aspects,” Journal of Infection and Public Health, 2020.

9. S. A. Hassan, F. N. Sheikh, S. Jamal, J. K. Ezeh, and A. Akhtar, “Coronavirus (covid-19): a review of clinical features, diagnosis, and treatment,” Cureus, vol. 12, no. 3, 2020.

10. A. Alimadadi, S. Aryal, I. Manandhar, P. B. Munroe, B. Joe, and X. Cheng, “Artificial intelligence and machine learning to fight covid-19,” 2020.

11. G. S. Randhawa, M. P. Soltysiak, H. El Roz, C. P. de Souza, K. A. Hill, and L. Kari, “Machine learning using intrinsic genomic signatures for rapid classification of novel pathogens: Covid-19 case study,” Plos one, vol. 15, no. 4, p. e0232391, 2020.

12. L. Huang, R. Han, T. Ai, P. Yu, H. Kang, Q. Tao, and L. Xia, “Serial quantitative chest ct assessment of covid-19: Deep-learning approach,” Radiology: Cardiothoracic Imaging, vol. 2, no. 2, p. e200075, 2020.

13. S. Minaee, R. Kafieh, M. Sonka, S. Yazdani, and G. J. Soufi, “Deep-covid: Predicting covid-19 from chest x-ray images using deep transfer learning,” *arXiv preprint arXiv:2004-09363*, 2020.

14. I. D. Apostolopoulos and T. A. Mpesiana, “Covid-19: automatic detection from x-ray images utilizing transfer learning with convolutional neural networks,” Physical and Engineering Sciences in Medicine, p. 1, 2020.

15. A. Borghesi and R. Maroldi, “Covid-19 outbreak in italy: experimental chest x-ray scoring system for quantifying and monitoring disease progression,” La radiologia medica, p. 1, 2020.

16. D. Das, K. Santosh, and U. Pal, “Truncated inception net: Covid-19 outbreak screening using chest x-rays,” Physical and Engineering Sciences in Medicine, pp. 1—11, 2020.

17. R. M. Pereira, D. Bertolini, L. O. Teixeira, C. N. Silla Jr, and Y. M. Costa, “Covid-19 identification in chest x-ray images on flat and hierarchical classification scenarios,” Computer Methods and Programs in Biomedicine, p. 105532, 2020.

18. T. Ozturk, M. Talo, E. A. Yildirim, U. B. Baloglu, O. Yildirim, and U. R. Acharya, “Automated detection of covid-19 cases using deep neural networks with x-ray images,” Computers in Biology and Medicine, p. 103792, 2020.

19. L. Wang and A. Wong, “Covid-net: A tailored deep convolutional neural network design for detection of covid-19 cases from chest x-ray images,” *arXiv preprint arXiv:2003.09871*, 2020.

20. A. Wong, M. J. Shafiee, B. Chwyl, and F. Li, “Ferminets: Learning generative machines to generate efficient neural networks via generative synthesis,” *arXiv preprint arXiv:1809.05989*, 2018.

21. A. Narin, C. Kaya, and Z. Pamuk, “Automatic detection of coronavirus disease (covid-19) using x-ray images and deep convolutional neural networks,” *arXiv preprint arXiv:2003.10849*, 2020.

22. J. P. Cohen, P. Morrison, and L. Dao, “Covid-19 image data collection,” *arXiv 2003.11597*, 2020. [Online]. Available: https://github.com/ieee8023/covid-chestxray-dataset

23. P. Mooney, “Chest x-ray images (pneumonia),” Mar 2018. [Online]. Available: https://www.kaggle.com/paultimothymooney/chest-xray-pneumonia

24. “U.s. national library of medicine. tuberculosis chest x-ray image data sets,” 2020. [Online]. Available: https:/https://ceb.nlm.nih.gov/tuberculosis-chest-X-rayimage-data-sets/

25. X. Wang, Y. Peng, L. Lu, Z. Lu, M. Bagheri, and R. M. Summers, “Chestx-ray8: Hospital-scale chest x-ray database and benchmarks on weakly-supervised classification and localization of common thorax diseases,” in Proceedings of the IEEE conference on computer vision and pattern recognition, 2017, pp. 2097—2106.

26. Z. Q. L. Linda Wang and A. Wong, “Covid-net: A tailored deep convolutional neural network design for detection of covid-19 cases from chest radiography images,” 2020.

27. M. E. Chowdhury, T. Rahman, A. Khandakar, R. Mazhar, M. A. Kadir, Z. B. Mahbub, K. R. Islam, M. S. Khan, A. Iqbal, N. Al-Emadi et al., “Can ai help in screening viral and covid-19 pneumonia?” *arXiv preprint arXiv:2003.13145*, 2020.

28. E. E.-D. Hemdan, M. A. Shouman, and M. E. Karar, “Covidx-net: A framework of deep learning classifiers to diagnose covid-19 in x-ray images,” *arXiv preprint arXiv:2003.11055*, 2020.

29. J. Zhang, Y. Xie, Y. Li, C. Shen, and Y. Xia, “Covid-19 screening on chest x-ray images using deep learning based anomaly detection,” *arXiv preprint arXiv:2003.12338*, 2020.

30. Y. Bai, L. Yao, T. Wei, F. Tian, D.-Y. Jin, L. Chen, and M. Wang, “Presumed asymptomatic carrier transmission of covid-19,” Jama, vol. 323, no. 14, pp. 1406—1407, 2020.

31. A. Abbas, M. M. Abdelsamea, and M. M. Gaber, “Classification of covid-19 in chest x-ray images using detrac deep convolutional neural network,” *arXiv preprint arXiv:2003.13815*, 2020.

32. H. S. Maghdid, A. T. Asaad, K. Z. Ghafoor, A. S. Sadiq, and M. K. Khan, “Diagnosing covid-19 pneumonia from x-ray and ct images using deep learning and transfer learning algorithms,” *arXiv preprint arXiv:2004.00038*, 2020.

33. P. Afshar, S. Heidarian, F. Naderkhani, A. Oikonomou, K. N. Plataniotis, and A. Mohammadi, “Covid-caps: A capsule network-based framework for identification of covid-19 cases from x-ray images,” *arXiv preprint arXiv:2004.02696*, 2020.

34. M. J. Horry, M. Paul, A. Ulhaq, B. Pradhan, M. Saha, N. Shukla et al., “X-ray image based covid-19 detection using pre-trained deep learning models,” 2020.

35. K. Sun, Y. Zhao, B. Jiang, T. Cheng, B. Xiao, D. Liu, Y. Mu, X. Wang, W. Liu, and J. Wang, “High-resolution representations for labeling pixels and regions,” *arXiv preprint arXiv:1904.04514*, 2019.

36. O. Ronneberger, P. Fischer, and T. Brox, “U-net: Convolutional networks for biomedical image segmentation,” in International Conference on Medical image computing and computer-assisted intervention. Springer, 2015, pp. 234–241.

37. J. Irvin, P. Rajpurkar, M. Ko, Y. Yu, S. Ciurea-Ilcus, C. Chute, H. Marklund, B. Haghgoo, R. Ball, K. Shpanskaya et al., “Chexpert: A large chest radiograph dataset with uncertainty labels and expert comparison,” in Proceedings of the AAAI Conference on Artificial Intelligence, vol. 33, 2019, pp. 590–597.

38. S. Jaeger, S. Candemir, S. Antani, Y.-X. J. Wang, P.-X. Lu, and G. Thoma, “Two public chest x-ray datasets for computer-aided screening of pulmonary diseases,” Quantitative imaging in medicine and surgery, vol. 4, no. 6, p. 475, 2014.

39. T. Zhang, G.-J. Qi, B. Xiao, and J. Wang, “Interleaved group convolutions,” in Proceedings of the IEEE international conference on computer vision, 2017, pp. 4373–4382.

40. F. Milletari, N. Navab, and S.-A. Ahmadi, “V-net: Fully convolutional neural networks for volumetric medical image segmentation,” in 2016 fourth international conference on 3D vision (3DV). IEEE, 2016, pp. 565–571.

